# Rapid turnaround multiplex sequencing of SARS-CoV-2: comparing tiling amplicon protocol performance

**DOI:** 10.1101/2021.12.28.21268461

**Authors:** Bede Constantinides, Hermione Webster, Jessica Gentry, Jasmine Bastable, Laura Dunn, Sarah Oakley, Jeremy Swann, Nicholas D Sanderson, Philip W Fowler, Geoffrey Ma, Gillian Rodger, Lucinda Barrett, Katie Jeffery, Timothy EA Peto, Nicole Stoesser, Teresa Street, Derrick W Crook

## Abstract

Genome sequencing is pivotal to virus surveillance, revealing the emergence and global dissemination of acquired genetic mutations. Amplicon sequencing has proven effective for sequencing SARS-CoV-2, but prevalent mutations disrupting primer binding sites have necessitated frequent revision of sequencing protocols in order to maintain performance for emerging virus lineages. We compared the performance of Oxford Nanopore Technologies (ONT) Midnight and ARTIC tiling amplicon protocols using 96-plexed nanopore sequencing of 193 Delta-lineage SARS-CoV-2 clinical specimens with complete paired results and 71 S-gene target failure-selected specimens collected during emergence of Omicron BA.1. For Delta samples, ARTIC v4 recovered ≥90% of the genome in 82.4% (159/193) of specimens compared with 71.5% (138/193) for Midnight. ARTIC v4 was more robust in low-titre, high-Ct specimens, whereas Midnight achieved slightly greater completeness at lower Ct. In the S-gene target failure-selected cohort, revised Midnight v2 and ARTIC v4.1 primer schemes substantially improved genome completeness: ≥99% genome recovery was achieved in 74.6% (53/71) with Midnight v2 versus 18.3% (13/71) with Midnight, and 57.7% (41/71) with ARTIC v4.1 versus 0% (0/71) with ARTIC v4. Estimated consumable costs were lower for Midnight than ARTIC (£12 versus £16 per sample at Q1 2022 prices), and the interval from reverse transcription to flow cell loading was shorter (~7 versus ~9.5 hours). Midnight proved a practical alternative for rapid nanopore sequencing, with lower consumable costs and shorter preparation time, although ARTIC v4 was more robust in high-Ct Delta specimens.

**Impact statement:** Tiling amplicon whole genome sequencing enables fast and cost-effective virus sequencing, yet is adversely affected by mutations at primer binding sites. We evaluated ARTIC and Midnight nanopore sequencing protocols using SARS-CoV-2 clinical samples in terms of genome recovery, turnaround time and consumable costs. Our evaluation contributed to deployment of a nanopore sequencing service embedded in an NHS hospital microbiology laboratory supporting genomic surveillance and subsequently the RECOVERY platform trial.

## Introduction

SARS-CoV-2 sequencing is pivotal to global surveillance and the international and national public health response. The ARTIC Network’s open source tiled amplicon protocol is widely used for sequencing SARS-CoV-2, has been updated to reflect changes in circulating lineages internationally, and is suitable for low cost, highly-multiplexed sequencing using popular Illumina and Oxford Nanopore Technologies (ONT) platforms (Tyson et al., 2020; Pipelines R&D et al., 2022; Quick & Lansdowne, 2024). ONT’s Midnight protocol is a commercially available nanopore alternative using 1200-bp tiled amplicons and rapid barcoding, offering reduced turnaround time and cost relative to ligation-based nanopore library preparation (Freed et al., 2020). Subsequent evaluations have further demonstrated the value of tiling amplicon approaches for SARS-CoV-2 sequencing, highlighting tradeoffs between genome completeness, susceptibility to primer dropout, cost, and turnaround time (Bayega et al., 2025; Koskela Von Sydow et al., 2023). We sought to compare the performance of ARTIC and Midnight protocols to support the establishment of a routine SARS-CoV-2 sequencing service in an NHS clinical laboratory. For Omicron (BA.1) lineages we also evaluated revised prototype versions of both tiling amplicon protocols, to which we refer as Midnight v2 and ARTIC v4.1 (Freed, 2021; Quick, 2022). Reflecting prevailing lineages in the United Kingdom in late 2021, we sequenced clinical samples comprising the Delta-associated Pango lineages AY.4, AY.4.2 and B.1.617.2, and the Omicron BA.1-like lineage as assigned by Pangolin (Rambaut et al., 2020; O’Toole et al., 2021).

## Results

During genomic surveillance of patients and health care workers, we characterised 198 distinct PCR-positive clinical specimens of Delta lineage collected during November 2021 and 71 S-gene target failure (SGTF)-selected specimens collected in December 2021 comprising mostly Omicron samples. Among the latter, 48 were assigned Omicron by Pangolin’s rule-based Scorpio classifier. Two Delta specimens were excluded from the primary analysis as suspected ARTIC workflow failures. Of the remaining 196 specimens, 193 (98.5%) had complete paired results for Midnight and ARTIC v4 and formed the basis of this comparison.

For Delta samples, using a criterion of at least 90% coverage at depth ≥20, ARTIC v4 recovered the greatest proportion of nearly-complete genomes (82.4%; 159/193), followed by Midnight (71.5%; 138/193), and ARTIC v3 (34.1%; 14/41). Among the 193 paired specimens, 22 reached the threshold only with ARTIC v4 and one only with Midnight (two-sided exact McNemar p<0.00001). Including the two suspected ARTIC workflow failures gave ≥90% genome recovery in 159/195 (81.5%) specimens with ARTIC v4 and 140/195 (71.8%) with Midnight. Among 193 specimens with complete paired results, mean cov20 was higher with ARTIC v4 than with Midnight (91.4% versus 79.3%), whereas median cov20 was slightly higher with Midnight (99.4% versus 98.2%). This difference reflects ARTIC v4’s greater recovery in high-Ct specimens: Midnight yielded higher cov20 in 121 specimens (62.7%), ARTIC v4 in 71 (36.8%), and the protocols were tied in one specimen. ARTIC v3, evaluated separately in 41 specimens from sample set 1, had mean and median cov20 values of 75.4% and 82.5%, respectively. At Ct <25, Midnight had slightly higher median cov20 than ARTIC v4 (99.4% versus 98.8%), although its mean cov20 was lower (96.9% versus 98.5%) because of several poor results. At Ct ≥25, mean cov20 was 45.5% with Midnight and 77.8% with ARTIC v4 (Figure 1A; Ct distributions are shown in Figure S2). Genome recovery (cov20) and unambiguous consensus sequence coverage were strongly correlated (Pearson’s *r* > 0.9999, p<0.00001; Figure S3). ARTIC v4 notably suffered from a ubiquitous dropout at 6850-7080bp (ORF1a) in our testing (Figure 2).

**Fig 1.**
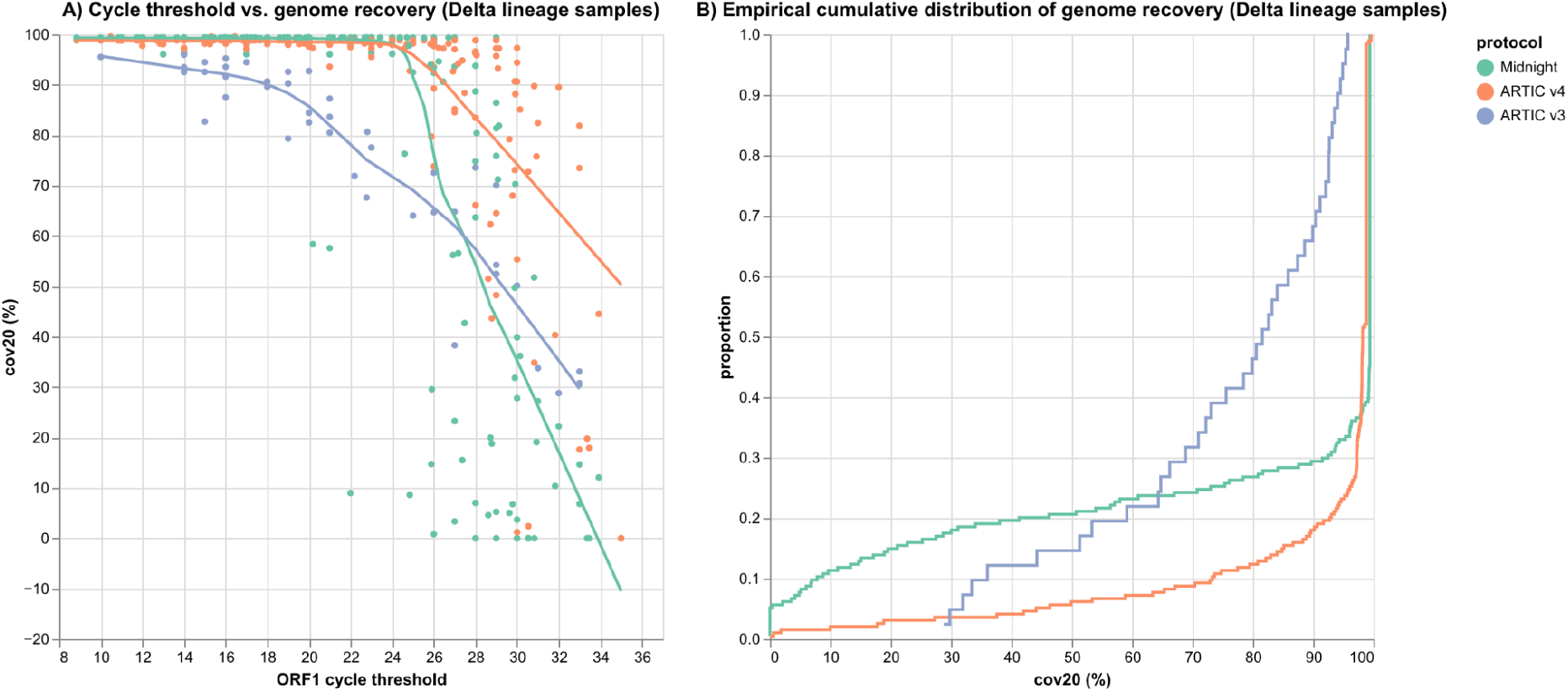
A) TaqPath ORF1 gene cycle threshold (Ct) vs. genome recovery at depth ≥20 (cov20) in 193 Delta lineage samples with complete paired Midnight and ARTIC v4 results, drawn from the 196-sample cohort, with LOESS regression lines. B) Empirical cumulative distribution of cov20. ARTIC v4 retained higher genome recovery as Ct increased. At lower Ct, Midnight generally had marginally higher completeness, although ARTIC v4 had fewer severe failures. Shown are sample sets 1-3 for Midnight and ARTIC v4, and sample set 1 only for ARTIC v3, which performed poorly.

**Fig 2.**
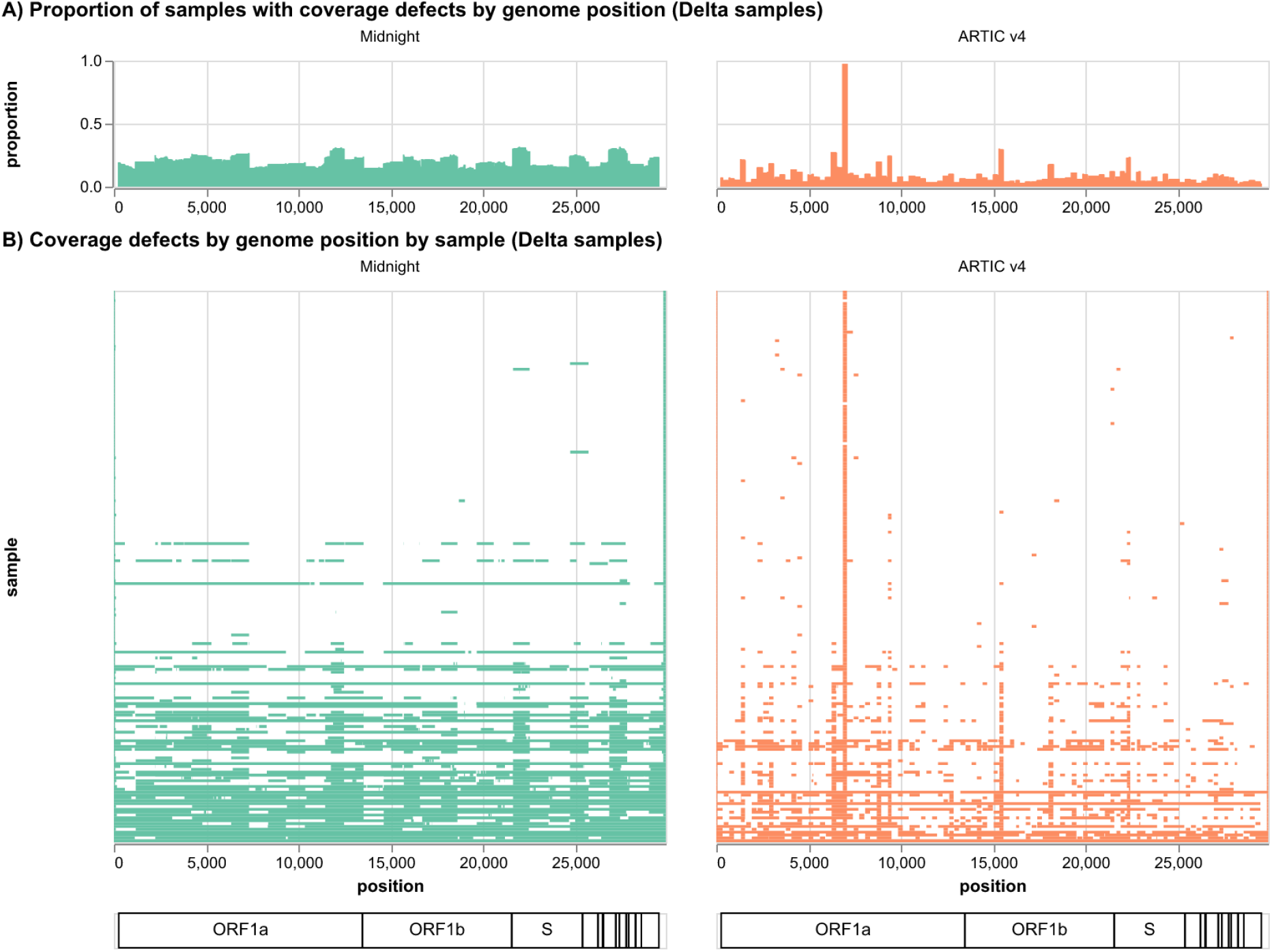
Coverage defect maps of Midnight and ARTIC v4 protocols applied to 193 Delta lineage clinical samples with complete paired results (sample sets 1-3). A) Proportion of samples with coverage defects (depth <20) by reference genome position. B) Per-sample distribution of coverage defects, sorted by ascending ORF1ab Ct. ARTIC v4 exhibited a systematic defect at approximately 6850-7080bp (ORF1a) (right).

For Omicron samples, we compared performance of the then-commercially available Midnight and ARTIC v4 protocols alongside revised Midnight v2 and ARTIC v4.1 protocols, both featuring revised primer mixtures designed to address defects associated with the constellation of mutations defining Omicron lineage SARS-CoV-2. The revised protocols both offered improved performance compared to respective previous versions, and exhibited similar overall performance to one another in the test panel of SGTF-selected clinical Omicron samples. Using the same criterion of ≥90% cov20, ARTIC v4.1 and v4 recovered the same proportion of nearly-complete Omicron genomes (88.7%; 63/71), followed by Midnight v2 (85.9%; 61/71), and Midnight (83.1%; 59/71). Using a more stringent measure of genome recovery (≥99% cov20) highlighted much greater differences between protocols: Midnight v2 recovered 74.6% (53/71) of samples, followed by ARTIC v4.1 (57.7%; 41/71), followed by Midnight (18.3%; 13/71), followed by ARTIC v4 (0%; 0/71). See Figure 3 for the empirical distribution of cov20 by protocol. Giving rise to the revised protocols’ improved performance were reductions in amplicon failure with respect to existing versions. A dropout in Midnight protocol affecting genes ORF8, ORF9b and N appeared fully resolved in Midnight v2, as were ARTIC v4 dropouts in S gene (Figure 4). Dropouts in ARTIC v4’s E and M genes appeared largely if not entirely addressed by the v4.1 protocol (Figure 4), although it should be noted that the V4.1 primers used here were unbalanced, and that balancing would be expected to further improve performance.

**Fig 3.**
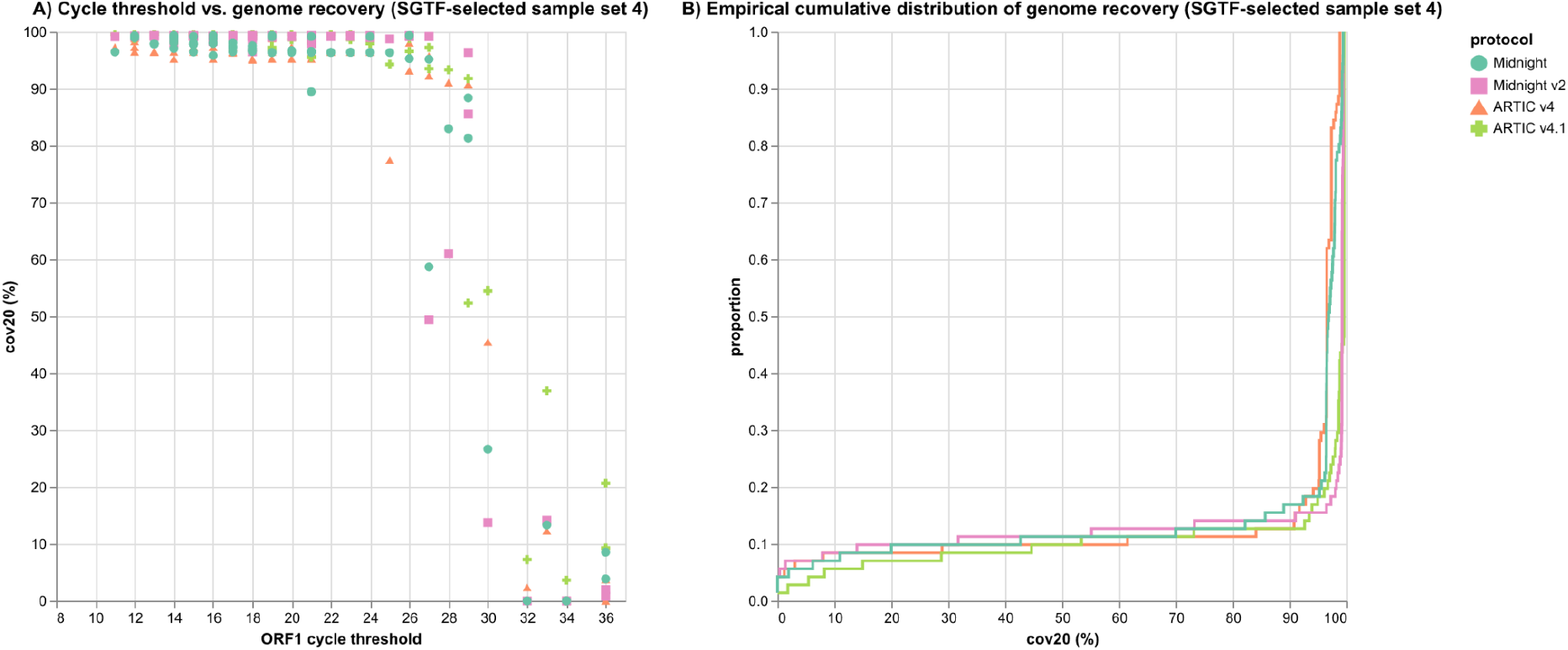
A) Comparison of existing and revised Midnight and ARTIC protocols in terms of ORF1ab Ct vs. genome recovery (cov20) in the 71 SGTF-selected samples in sample set 4, of which 48 were assigned to Omicron BA.1. LOESS lines omitted for lack of high Ct samples. B) Cumulative empirical cov20 distributions for Midnight, Midnight v2, ARTIC v4 and ARTIC v4.1.

**Fig 4.**
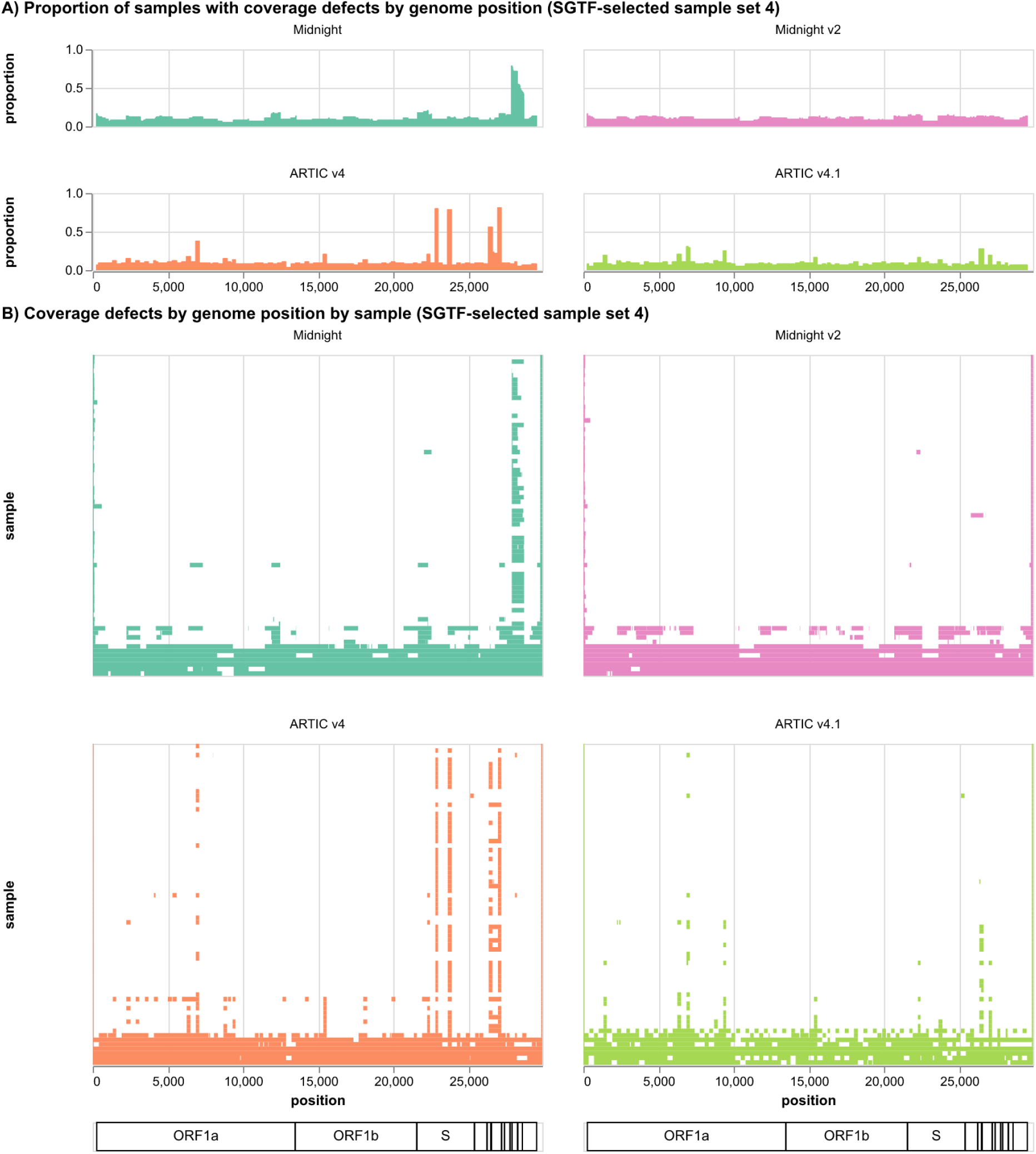
Coverage defect maps of existing and revised Midnight, ARTIC protocols applied to 71 SGTF-selected clinical samples in sample set 4, of which 48 were assigned Omicron BA.1. A) Proportion of samples with coverage defects (depth <20) by reference position. B) Per-sample distribution of coverage defects, sorted by ascending ORF1ab Ct. The additional primer proposed by Freed (2021) and used in the Midnight v2 protocol addresses the amplicon failure at ~28kb associated with the v1 commercial kit. For ARTIC v4.1, several new primers, some with revised coordinates, largely address the amplicon failures associated with Omicron BA.1 when using the v4 primers.

## Discussion

Community-developed open source amplicon sequencing protocols have seen widespread adoption in the SARS-CoV-2 pandemic, enabling reliable and cost-effective whole genome sequencing, maximising the global reach of genomic surveillance. Because amplicon sequencing performance is sensitive to mutations at primer binding sites, tiling amplicon schemes designed against early SARS-CoV-2 genomes have required repeated revision as viral diversity accumulated. Subsequent benchmarking has confirmed protocol-associated differences in genome completeness and primer dropout, including between ARTIC and longer amplicon protocols such as Midnight (Bayega et al., 2025).

Both protocols performed well in low-Ct Delta samples. Midnight often achieved slightly greater completeness, but ARTIC v4 had fewer severe failures and was more robust at high Ct (Figure 1A). In the SGTF-selected, predominantly Omicron BA.1 sample set, both existing protocols showed reduced performance, with a marked effect on ARTIC v4 through dropouts in the gene encoding spike protein. The revised Midnight v2 and ARTIC v4.1 protocols evaluated here both mitigated these issues and showed similar recovery across the predominantly low-Ct sample set, although dropouts occurred in different genomic regions. Neither revised protocol was evaluated here in its final commercial form, and the ARTIC v4.1 primer pools were unbalanced, likely to its detriment as tested. Using transposase-based rapid barcoding rather than the ligation-based native barcoding workflow used by ARTIC, Midnight had lower consumable costs and a shorter estimated interval from reverse transcription to flow cell loading of approximately 7 hours versus 9.5 hours. A subsequent independent comparison likewise found ARTIC and Midnight to be robust and scalable approaches for SARS-CoV-2 sequencing, with shorter library preparation for the Midnight workflow (Koskela Von Sydow et al., 2023). Our analysis did not evaluate demultiplexing accuracy; whereas Midnight used rapid barcoding, the ARTIC protocol used here attached barcodes to both ends of fragments, permitting more stringent barcode assignment. Together, these findings support the Midnight protocol as a practical alternative to ARTIC for rapid nanopore SARS-CoV-2 sequencing, with shorter library preparation and lower consumable costs albeit reduced genome recovery in low-titre, high-Ct samples.

The evaluation also contributed to the deployment of a DHSC Pillar 1 Surge sequencing service within an NHS clinical microbiology laboratory in an Oxfordshire teaching hospital. This sequencing capability subsequently supported the RECOVERY platform trial, including sequencing of samples for the assessment of mutations associated with resistance to molnupiravir or nirmatrelvir–ritonavir (RECOVERY Collaborative Group, 2025).

## Methods

### Clinical samples

Clinical samples used in this evaluation were laboratory discards obtained after routine diagnostic workflows were complete. Sample sets 1–3 contained 198 distinct PCR-positive Delta-lineage specimens. Two were excluded from the primary analyses as suspected ARTIC workflow failures. Both recovered more than 95% of the genome with Midnight but less than 2% with ARTIC v4. This left 196 specimens, of which 193 had complete paired Midnight and ARTIC v4 results. ARTIC v3 was evaluated separately in 41 specimens from sample set 1. One specimen appeared in both sample sets 1 and 3; for this specimen, the sample set 1 results were used. Clinical negative controls were included throughout each plate.

### RNA extraction

RNA was extracted from nasopharyngeal swabs using the KingFisher Flex platform and MagMax™ 96 Viral/Pathogen Nucleic Acid Isolation kit (Thermo Fisher Scientific, Waltham, MA, USA), following the 400µl extraction protocol. To achieve a higher yield of RNA for sequencing set 4 (predominantly Omicron samples), RNA was extracted from two 400µl sample inputs eluted into 100µl each which were combined prior to RT-PCR. RT-PCR, performed on a QuantStudio 5 with the TaqPath COVID-19 RT-PCR kit (Thermo Fisher Scientific), identified positive SARS-CoV-2 samples based on N gene, S gene and ORF1ab amplification, as well as providing cycle threshold (Ct) values.

### Reverse Transcription

Reverse transcription (RT) conditions were identical for both ARTIC and Midnight sequencing protocols. A 4:1 volume ratio of RNA to LunaScript RT SuperMix (New England Biolabs, Ipswich, MA, USA) was incubated at 25°C for 2 minutes, 55°C 10 mins and 95°C for 1 minute. Each sample set included a negative control at this step.

### ARTIC protocols

After reverse transcription, sequencing was performed based on the ARTIC protocol (Quick & Lansdowne, 2024) using the NEBNext E7660 ARTIC SARS-CoV-2 Companion Kit for Oxford Nanopore Technologies (New England Biolabs) and following the Express Protocol. ARTIC v3 and v4 primers were used at a concentration of 10µM. For ARTIC v4.1, 10µM v4 primers were spiked with additional (v4.1) primer pools at 10µM. The ARTIC v3 protocol was evaluated only on sample set one (41 positive samples), and, performing relatively poorly, was not evaluated further.

### Midnight protocol

The Midnight protocol (Freed et al., 2020), was followed using the Midnight RT PCR Expansion (SQK-RBK110.96 and EXP-MRT001) (ONT, Oxford, UK). PCR conditions differed between sample sets, reflecting changing manufacturer best practices. For sample set one, the Midnight revision H protocol was followed, with PCR annealing and extension conditions set at 65°C for 5 minutes. For sample sets two, three and four, protocol revision M was followed, with PCR annealing and extension conditions respectively set at 61°C for 2 minutes and 65°C for 3 minutes. Revised Midnight v2 (MRT001.20) primers were used for sample set 4 to address amplicon dropout in Omicron (BA.1).

### Sequencing

Pooled libraries generated by both protocols were sequenced with an ONT GridION using a total of eleven ONT R9.4.1 flow cells. Consumable costs were estimated at £12 per sample for Midnight and £16 for ARTIC based on 96-plexing consumables at Q1 2022 prices. Estimated elapsed times from reverse transcription to flow cell loading were approximately 7 hours for Midnight and 9.5 hours for ARTIC.

### Sequence analysis

Sequences were basecalled and demultiplexed using Guppy 5.0.16 in high accuracy mode. For ARTIC libraries, barcode sequences were required at both ends of each read, as appropriate for the ligation sequencing kit used. For Midnight libraries prepared using rapid barcoding, a barcode at one end of each read was required for assignment to a barcode. Sequence analysis was performed using a forked version of the EPI2ME Labs wf-artic Nextflow pipeline (https://github.com/bede/wf-artic/tree/0.3.6-patched-fast), namely incorporating ARTIC Fieldbioinformatics (Loman, 2020) and Pangolin (O’Toole et al., 2021). Depth of coverage and cov20 were calculated from primer-trimmed BAM files using Samtools (Li et al., 2009) depth, including aligned deletions in the depth calculation. Pangolin version 3.1.17 with PangoLEARN version 2021-12-06 was used. JupyterLab (Kluyver et al., 2016) was used for further analysis and visualisation. Paired Delta success rates (cov20 ≥90%) were compared using a two-sided exact McNemar test.

### Data summary

Sequencing reads are publicly deposited in ENA BioProject PRJEB124656. Human sequences were depleted from deposited reads using Deacon 0.17.0 (Constantinides et al., 2025) with default parameters. Figures can be reproduced using the code notebook in the permanently archived supporting GitHub repository (Bede Constantinides, 2026), from either the committed genome assemblies, or by downloading FASTQ reads using the helper script. This repository contains relevant metadata including cycle thresholds and anonymised laboratory identifiers. Sequences of viral RNA extracted from cultured samples used in this study are also deposited under ENA BioProject PRJEB50520 (Constantinides et al., 2024).

### Ethical approval

Sample processing and analysis were covered by ethical approval for the use of anonymised oro- or nasopharyngeal specimens from patients for the diagnosis of influenza and other respiratory pathogens, including SARS-CoV-2 (North West–Greater Manchester South Research Ethics Committee [REC], REC Ref: 19/NW/0730).

## Data Availability

Sequencing reads are publicly deposited in ENA BioProject PRJEB124656. Figures can be reproduced using the code notebook in the Zenodo-archived supporting GitHub repository https://github.com/bede/sars2-midnight-eval.

https://zenodo.org/records/22260506

https://www.ebi.ac.uk/ena/browser/view/PRJEB124656

## Acknowledgements

We thank Joshua Quick for developing and sharing prototype ARTIC v4.1 primers for testing. Large language models were used in editing the second revision of this manuscript.

## Author contributions

Bede Constantinides: Investigation, Formal analysis, Visualisation, Validation, Software, Data Curation, Resources, Writing - Original Draft. Hermione Webster: Investigation, Data Curation, Resources, Writing - Original Draft. Jessica Gentry: Investigation. Jasmine Bastable: Investigation. Laura Dunn: Investigation. Sarah Oakley: Investigation. Jeremy Swann: Resources. Nicholas Sanderson: Resources. Philip W Fowler: Writing - Review & Editing. Geoffrey Ma: Resources. Gillian Rodger: Investigation. Lucinda Barrett: Resources. Katie Jeffery: Resources. Timothy EA Peto: Writing - Review & Editing. Nicole Stoesser: Funding acquisition, Writing - Review & Editing. Teresa Street: Supervision, Methodology, Conceptualisation, Writing - Original Draft. Derrick W Crook: Supervision, Funding acquisition, Conceptualisation, Writing - Review & Editing.

## Supplementary figures

**S. fig. 1.**
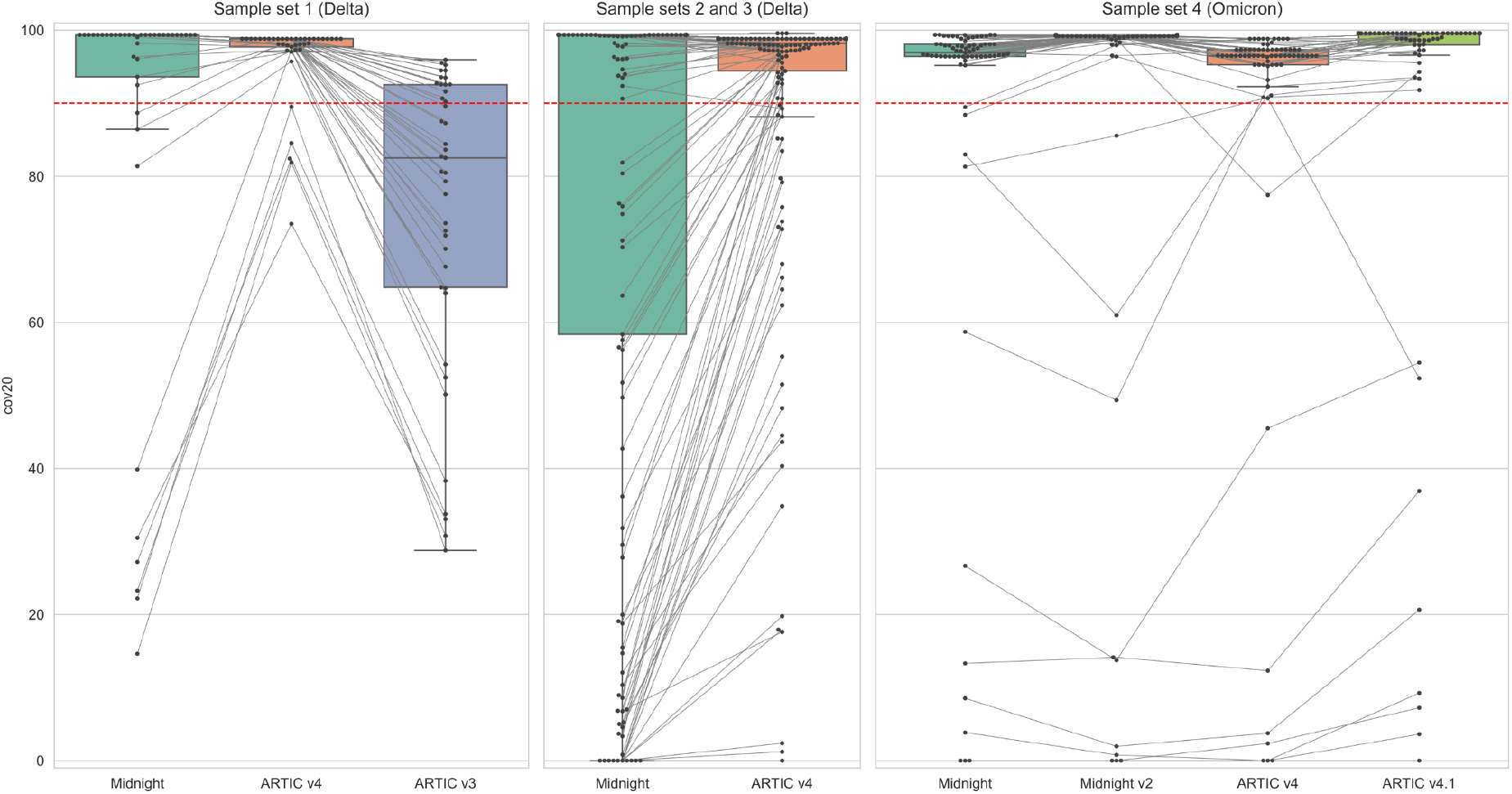
Genome recovery at depth ≥20 in head-to-head sequencing of 264 SARS-CoV-2 clinical samples, comprising 193 Delta lineage samples with complete paired results and 71 SGTF-selected samples, over a total of 11 runs. Red lines correspond to a widely used lower bound on genome recovery for lineage assignment (90%).

**S. fig. 2.**
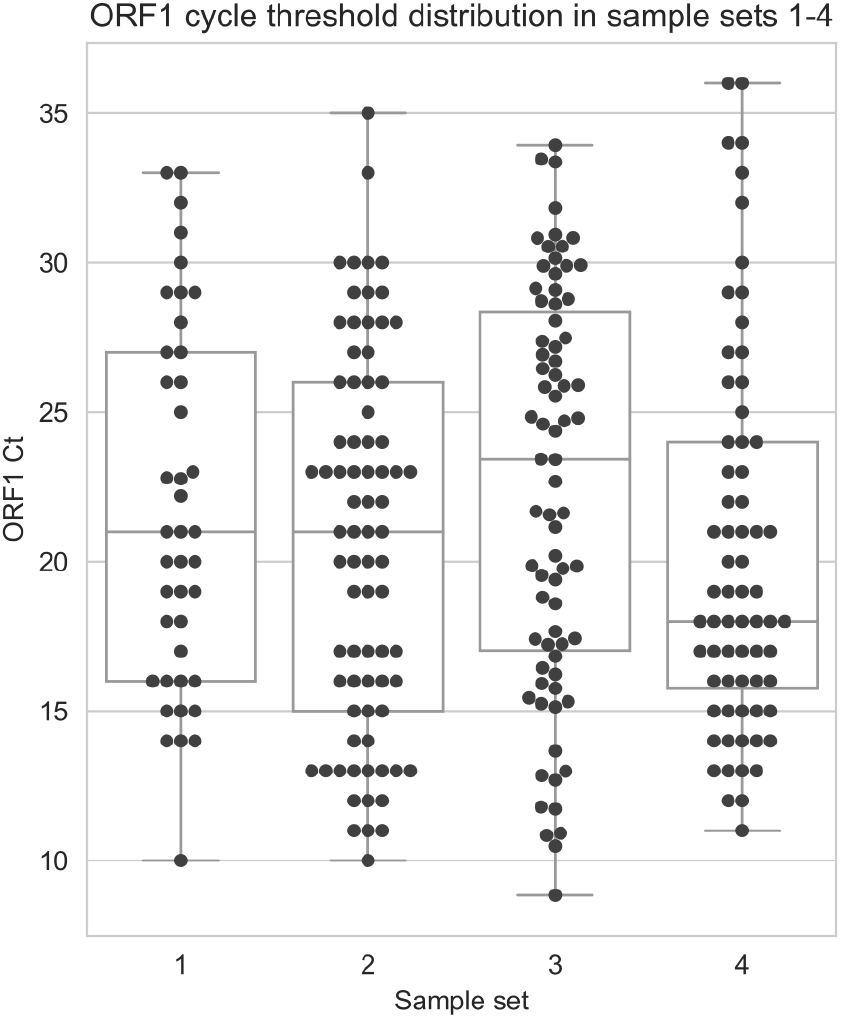
TaqPath RT-PCR ORF1ab Ct distribution for sample sets 1-4.

**S. fig. 3.**
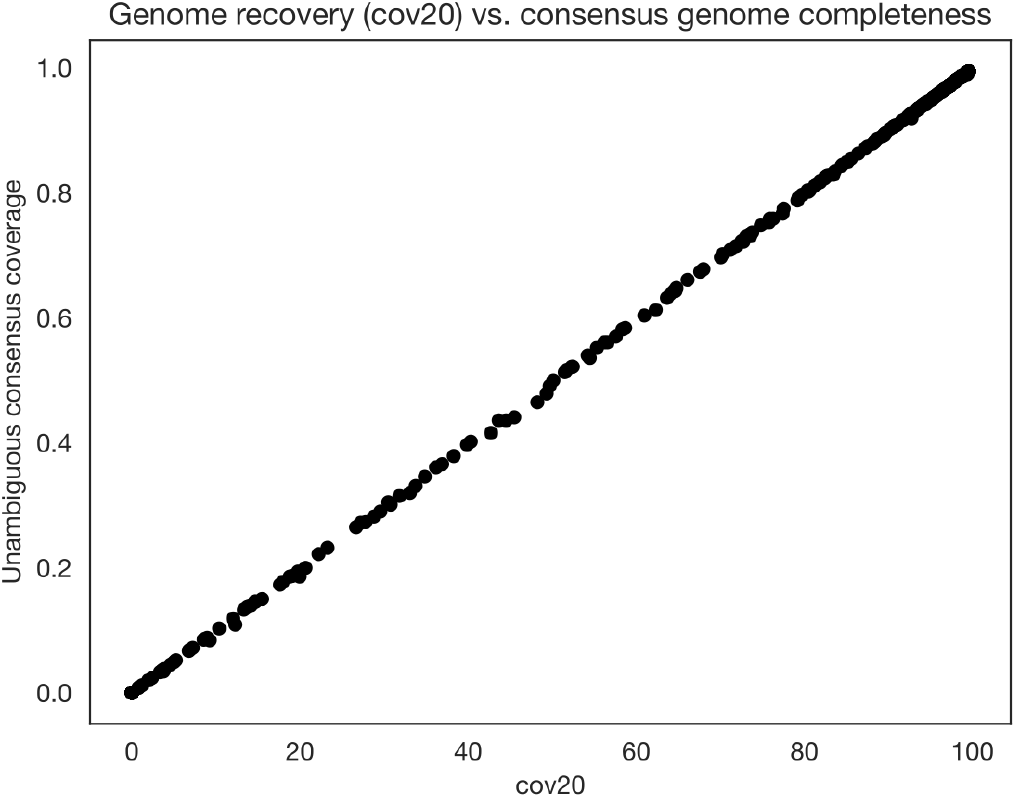
Genome recovery vs. unambiguous consensus genome coverage. Pearson r = 0.99998, p<0.00001.

## References

Bayega, A., Reiling, S. J., Liu, J. L., Dubuc, I., Gravel, A., Flamand, L., & Ragoussis, J. (2025). A Benchmark of methods for SARS-CoV-2 whole genome sequencing and development of a more sensitive method. Frontiers in Genetics, 16, 1516791. 10.3389/fgene.2025.1516791

Bede Constantinides. (2026). bede/sars2-midnight-eval: 0.1.0 (Version 0.1.0) [Computer software]. Zenodo. 10.5281/ZENODO.22260506

Constantinides, B., Lees, J., & Crook, D. W. (2025). Deacon: Fast sequence filtering andcontaminant depletion. 10.1101/2025.06.09.658732

Constantinides, B., Webster, H., Rodger, G., Hunt, M., Supasa, P., Dejnirattisai, W., Liu, C., Mongkolsapaya, J., Screaton, G. R., & Crook, D. W. (2024, February). A diverse reference set of cultured SARS-CoV-2 genomes sequenced using various amplification methods and instrument platforms. BioStudies, EMBL-EBI.10.6019/S-BSST1334

Freed, N. E., Vlková, M., Faisal, M. B., & Silander, O. K. (2020). Rapid and inexpensive whole-genome sequencing of SARS-CoV-2 using 1200 bp tiled amplicons and Oxford Nanopore Rapid Barcoding. Biology Methods and Protocols, 5(1), bpaa014. 10.1093/biomethods/bpaa014

Kluyver, T., Ragan-Kelley, B., Pérez, F., Granger, B., Bussonnier, M., Frederic, J., Kelley, K., Hamrick, J., Grout, J., Corlay, S., Ivanov, P., Avila, D., Abdalla, S., & Willing, C. (2016). Jupyter Notebooks – a publishing format for reproducible computational workflows. In F. Loizides & B. Schmidt (Eds), Positioning and Power in Academic Publishing: Players, Agents and Agendas (pp. 87–90). IOS Press.

Koskela Von Sydow, A., Lindqvist, C. M., Asghar, N., Johansson, M., Sundqvist, M., Mölling, P., & Stenmark, B. (2023). Comparison of SARS-CoV-2 whole genome sequencing using tiled amplicon enrichment and bait hybridization. Scientific Reports, 13(1), 6461. 10.1038/s41598-023-33168-1

Li, H., Handsaker, B., Wysoker, A., Fennell, T., Ruan, J., Homer, N., Marth, G., Abecasis, G., Durbin, R., & 1000 Genome Project Data Processing Subgroup. (2009). The Sequence Alignment/Map format and SAMtools. Bioinformatics, 25(16), 2078–2079. 10.1093/bioinformatics/btp352

Loman, N. J. (2020). The ARTIC field bioinformatics pipeline [Computer software]. https://github.com/artic-network/fieldbioinformatics

Nikki Freed. (2021, November 27). In this case, you could order this primer: SARSCoV_1200_28_LEFT_27837T: TTTGTGCTTTTTAGCCTTTCTGtT (Original primer SARSCoV_1200_28_LEFT: TTTGTGCTTTTTAGCCTTTCTGcT) and add it at an equimolar amount to pool 2. We have not tested this exact correction though. [Tweet]. Twitter. https://twitter.com/freed_nikki/status/1464477522448433156

O’Toole, Á., Scher, E., Underwood, A., Jackson, B., Hill, V., McCrone, J. T., Colquhoun, R., Ruis, C., Abu-Dahab, K., Taylor, B., Yeats, C., Du Plessis, L., Maloney, D., Medd, N., Attwood, S. W., Aanensen, D. M., Holmes, E. C., Pybus, O. G., & Rambaut, A. (2021). Assignment of epidemiological lineages in an emerging pandemic using the pangolin tool. Virus Evolution, 7(2), veab064. 10.1093/ve/veab064

Pipelines R&D, D., Farr, B., Rajan, D., Dawson, E., Shirley, L., Quail, M., Park, N., Redshaw,N. F Bronner, I., Aigrain, L., Goodwin, S., Thurston, S., Lensing, S., Bonfield, J., James, K., Salmon, N., Beaver, C., Nelson, R. K. Jackson, D., … Johnston, I. (2022).COVID-19 ARTIC v4.1 Illumina library construction and sequencing protocol—Tailed method v2. 10.17504/protocols.io.j8nlk4b36g5r/v2

Quick, J. (2022, February 11). Artic-ncov2019/primer_schemes/nCoV-2019/V4.1 at master · artic-network/artic-ncov2019. https://github.com/artic-network/artic-ncov2019/tree/master/primer_schemes/nCoV-2019/V4.1

Quick, J., & Lansdowne, L. (2024). ARTIC SARS-CoV-2 sequencing protocol v4 (LSK114) v2. 10.17504/protocols.io.bp2l6n26rgqe/v4

Rambaut, A., Holmes, E. C., O’Toole, Á., Hill, V., McCrone, J. T., Ruis, C., du Plessis, L., & Pybus, O. G. (2020). A dynamic nomenclature proposal for SARS-CoV-2 lineages to assist genomic epidemiology. Nature Microbiology, 5(11), 1403–1407. 10.1038/s41564-020-0770-5

RECOVERY Collaborative Group. (2025). Molnupiravir or nirmatrelvir–ritonavir plus usual care versus usual care alone in patients admitted to hospital with COVID-19 (RECOVERY): A randomised, controlled, open-label, platform trial. The Lancet Infectious Diseases, 25(9), 1000–1010. 10.1016/S1473-3099(25)00093-3

Tyson, J. R., James, P., Stoddart, D., Sparks, N., Wickenhagen, A., Hall, G., Choi, J. H., Lapointe, H., Kamelian, K., Smith, A. D., Prystajecky, N., Goodfellow, I., Wilson, S. J., Harrigan, R., Snutch, T. P., Loman, N. J., & Quick, J. (2020). Improvements to the ARTIC multiplex PCR method for SARS-CoV-2 genome sequencing using nanopore. Genomics. 10.1101/2020.09.04.283077

